# Estimating effects of serum vitamin B12 levels on psychiatric disorders and cognitive impairment: a Mendelian randomization study

**DOI:** 10.1101/2024.01.23.24301678

**Authors:** Tianyuan Lu, Andrew D. Paterson

## Abstract

Vitamin B12 deficiency can lead to pernicious anemia, various neuropsychiatric diseases, and cognitive decline. However, it is unclear whether increasing vitamin B12 levels can help to prevent the onset of psychiatric disorders and cognitive impairment in the general population. Leveraging large-scale genome-wide association studies (GWASs), we conducted Mendelian randomization (MR) analyses to estimate the potential effects of serum vitamin B12 levels on eight psychiatric disorders, as well as educational attainment and cognitive performance. We performed sensitivity analyses by excluding genetic instruments that demonstrated potential horizontal pleiotropy. We conducted additional MR analyses utilizing within-sibship studies for depressive symptoms, educational attainment, and cognitive performance to mitigate bias due to potential residual confounding in GWASs. As a positive control, we confirmed that a one standard deviation increase in genetically increased vitamin B12 levels was strongly associated with a decreased odds of developing pernicious anemia (odds ratio, OR = 0.24; 95% confidence interval, CI: 0.15-0.40; p-value = 2.1x10^-8^). In contrast, MR estimates of vitamin B12 effects on all eight psychiatric disorders, educational attainment and cognitive performance largely overlapped with the null. In particular, based on the three most well-powered GWASs, a one standard deviation increase in genetically predicted vitamin B12 levels was associated with an OR of 1.01 for depression (95% CI: 0.97-1.04; p-value = 0.74), a 7.7x10^-3^ standard deviation increase in educational attainment (95% CI: -1.0x10^-2^-2.5x10^-2^; p-value = 0.39) and a 1.3x10^-2^ standard deviation increase in cognitive performance (95% CI: -8.8x10^-3^-3.5x10^-2^; p-value = 0.24). No significant associations between genetically predicted vitamin B12 levels and any of the outcomes were identified in sensitivity analyses excluding pleiotropic genetic instruments or MR analyses based on within-sibship studies. In summary, our findings suggest that increasing serum vitamin B12 levels may not protect against the investigated psychiatric disorders or cognitive impairment in the general population.

## Introduction

Vitamin B12, also known as cobalamin, is an important water-soluble vitamin^1^. Together with folate, vitamin B12 has multiple essential roles in various biological processes, such as DNA synthesis and methylation, maturation of erythrocytes, as well as fatty acid and amino acid metabolism^2–4^. Vitamin B12 also contributes to the maintenance of the nervous system via the synthesis of myelin and neuronal regeneration^5, 6^. For humans, vitamin B12 is obtained exclusively from dietary intake^1^.

Vitamin B12 deficiency affects a significant proportion of the global population, including both females and males and across all age groups^7, 8^. Vitamin B12 deficiency defined as serum vitamin B12 levels < 148 pmol per litre has been estimated to affect 2.9% of the adult population in the United States^9^. Untreated vitamin B12 deficiency can lead to pernicious anemia and can result in neurological manifestations, such as peripheral neuropathy, ataxia, various psychiatric disorders, as well as cognitive impairment^7, 8, 10^. Amongst individuals with depression or schizophrenia, randomized placebo-controlled trials have shown that folate plus vitamin B12 supplementation may preserve cognitive functions and delay the disease progression^11–13^. However, randomized placebo-controlled trials have not elucidated whether increasing vitamin B12 levels through supplementation can help to prevent the onset of psychiatric disorders and cognitive impairment in the general population^14^, where the majority does not have clinical vitamin B12 deficiency. Although a longitudinal study indicated that vitamin B12 levels are negatively associated with incident depressive symptoms in older adults^15^, these observational associations may be subject to confounding factors that are difficult to fully account for, such as socioeconomic status, lifestyle factors, and comorbidities.

Mendelian randomization (MR) provides an effective and inexpensive alternative for estimating potential effects of vitamin B12 levels in the general population. MR employs genetic variants as instruments for an exposure (i.e. vitamin B12 levels) and assesses the associations between the genetically predicted exposure and the outcomes under investigation^16, 17^. MR relies on three instrumental variable assumptions: (1) the genetic instruments should be associated with the exposure; (2) the genetic instruments should not be associated with any confounders; and (3) the genetic instruments should affect the outcome through the exposure and not through other mechanisms, which is known as the assumption of no horizontal pleiotropy. Given that genetic variants are typically randomized at conception, the first two MR assumptions are likely fulfilled by selecting variants significantly associated with serum B12 levels as genetic instruments from recent large-scale genome-wide association studies (GWASs)^18^, although the assumption of no horizontal pleiotropy requires further verification.

MR has been adopted to evaluate the relationships between serum vitamin B12 levels and several diseases and biomarkers. Suggestive evidence has indicated that increased vitamin B12 levels may marginally increase fasting glucose while reducing beta-cell function in the pancreas^19^. However, no explicit associations have been identified between vitamin B12 levels and coronary artery disease, type 2 diabetes, measures of obesity, or Alzheimer’s disease^19, 20^. Recently, a phenome-wide study based on the UK Biobank confirmed the association between increased vitamin B12 levels and reduced odds of pernicious anemia^21^. However, it remains unclear whether the genetically predicted vitamin B12 levels are associated with psychiatric disorders and cognitive impairment, due to limited statistical power in previous studies.

Therefore, in this study, we performed MR analyses to investigate the potential effects of serum vitamin B12 levels on eight common psychiatric disorders and cognitive impairment in the general population. To maximize statistical power, we utilized the GWASs with the largest sample sizes to date, encompassing attention-deficit/hyperactivity disorder (ADHD), anorexia nervosa, anxiety disorders, autism spectrum disorder, bipolar disorder, depression, obsessive-compulsive disorder (OCD), schizophrenia, as well as educational attainment and cognitive performance. Multiple sensitivity analyses were performed to address potential bias arising from violations of instrumental variable assumptions and residual confounding in GWASs. This study aims to contribute to the understanding of whether vitamin B12 supplementation could serve as a preventive measure against psychiatric disorders and cognitive impairment in the general population.

## Methods

### Genome-wide association studies of serum vitamin B12 and folate levels

Serum vitamin B12 levels were measured in three cohort studies^18^. GWAS meta-analyses were performed based on a total of 45,576 participants of European ancestry, including 37,283 individuals from an Icelandic cohort and 8,293 individuals from two Danish cohorts. Furthermore, GWAS meta-analyses for serum folate levels were performed in the same cohort studies, based on a total of 37,341 individuals. Vitamin B12 and folate levels were quantile normalized prior to analysis. Details of these GWAS meta-analyses have been described previously^18^. Independent genetic variants significantly associated with serum vitamin B12 and folate levels with a p-value < 2.2x10^-9^ (Bonferroni-corrected genome-wide significance threshold accounting for 22.9 million tested variants) were utilized as genetic instruments. We calculated the F-statistic of each genetic instrument, where an F-statistic > 10 would indicate a low risk of weak instrument bias^22^. The genetic instruments and the mapped protein-coding genes are summarized in **Supplementary Table S1**.

### Known genotype-phenotype associations involving genetic instruments

To assess the potential risk of horizontal pleiotropy, we evaluated whether the selected genetic instruments have been associated with other phenotypes. We queried each genetic instrument in the Open Targets Genetics database^23, 24^ (https://genetics.opentargets.org/, accessed December 15, 2023), which includes existing GWASs in the GWAS Catalog as well as GWASs conducted using the UK Biobank or FinnGen resources^25–28^. After excluding associations with vitamin B12 levels, folate levels, and pernicious anemia, we considered genetic instruments that are associated with one to five other phenotypes (p-value < 5.0x10^-8^) to be subject to a moderate risk of horizontal pleiotropy, and those associated with more than five other phenotypes to be subject to a high risk of horizontal pleiotropy. The following analyses focused on vitamin B12 levels because both of the two genetic instruments for folate levels demonstrated potential horizontal pleiotropy. All genotype-phenotype associations involving genetic instruments are summarized in **Supplementary Table S2**.

### Association between vitamin B12 levels and pernicious anemia

As a positive control, we assessed whether genetically predicted vitamin B12 and folate levels were associated with pernicious anemia. We obtained UK Biobank exome-based association statistics from the Genebass database^29^ (https://app.genebass.org/, accessed October 19, 2023) for all genetic instruments were available in the exome-sequencing data. Details regarding quality control, exome-side association studies, and data curation have been described previously^29^. We queried each genetic instrument for its associations with various types of anemia, based on International Classification of Diseases version 10 (ICD-10) codes or self-reported physician-made diagnoses.

We performed MR to assess the association between the genetically predicted vitamin B12 levels and each type of anemia using the weighted median method. This method can tolerate the inclusion of invalid genetic instruments and can provide consistent estimates with up to 50% of the information coming from invalid genetic instruments^30^. We additionally performed MR using the inverse variance weighted regression, penalized weighted median, simple mode, weighted mode, and MR-Egger method, respectively. A nominally significant MR-Egger intercept (p-value < 0.05) was considered evidence of directional pleiotropy^31^. We derived Wald ratio estimates to assess the association between the genetically predicted folate levels and each type of anemia, since only one genetic instrument for folate levels was available in the UK Biobank exome-sequencing data. MR analyses were conducted using the TwoSampleMR R package version 0.5.6^32^.

### Genome-wide association studies of psychiatric disorders, educational attainment, and cognitive performance

The summary statistics of the most recent large-scale GWAS meta-analyses for eight psychiatric disorders were obtained from the Psychiatric Genomics Consortium, including ADHD^33^ (N cases = 20,183, N controls = 35,191), anorexia nervosa^34^ (N cases = 16,992, N controls = 55,525), anxiety disorders^35^ (N cases = 7,016, N controls = 14,745), autism spectrum disorder^36^ (N cases = 18,381, N controls = 27,969), bipolar disorder^37^ (N cases = 41,917, N controls = 371,549), depression^38^ (N cases = 246,363, N controls = 561,190), OCD^39^ (N cases = 2,688, N controls = 7,037), and schizophrenia^40^ (N cases = 69,369, N controls = 236,642). The GWAS summary statistics for educational attainment^41^ (N = 765,283), measured as number of years of schooling completed, were obtained from the Social Science Genetic Association Consortium. The GWAS summary statistics for cognitive performance^42^ (N = 257,828), quantified based on various cognitive tests, were obtained from the Cognitive Genomics Consortium. Participants of these study cohorts were predominantly of European ancestry. Details of these GWASs are summarized in **Table 1**.

**Table 1.**
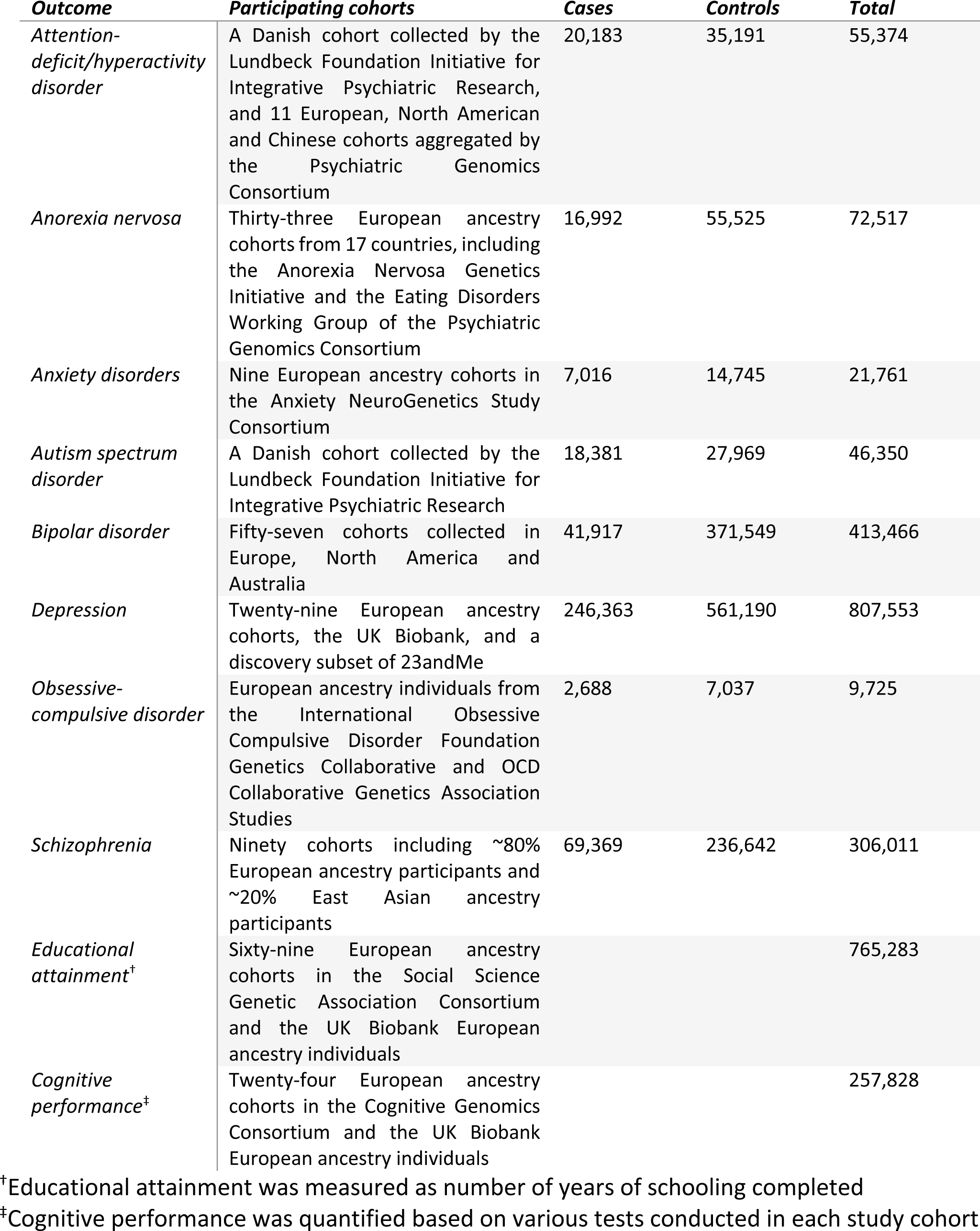
Genome-wide association studies of psychiatric disorders, educational attainment, and cognitive performance.

| **Outcome** | **Participating cohorts** | **Cases** | **Controls** | **Total** |
| --- | --- | --- | --- | --- |
| *Attention-deficit/hyperactivity disorder* | A Danish cohort collected by the Lundbeck Foundation Initiative for Integrative Psychiatric Research, and 11 European, North American and Chinese cohorts aggregated by the Psychiatric Genomics Consortium | 20,183 | 35,191 | 55,374 |
| *Anorexia nervosa* | Thirty-three European ancestry cohorts from 17 countries, including the Anorexia Nervosa Genetics Initiative and the Eating Disorders Working Group of the Psychiatric Genomics Consortium | 16,992 | 55,525 | 72,517 |
| *Anxiety disorders* | Nine European ancestry cohorts in the Anxiety NeuroGenetics Study Consortium | 7,016 | 14,745 | 21,761 |
| *Autism spectrum disorder* | A Danish cohort collected by the Lundbeck Foundation Initiative for Integrative Psychiatric Research | 18,381 | 27,969 | 46,350 |
| *Bipolar disorder* | Fifty-seven cohorts collected in Europe, North America and Australia | 41,917 | 371,549 | 413,466 |
| *Depression* | Twenty-nine European ancestry cohorts, the UK Biobank, and a discovery subset of 23andMe | 246,363 | 561,190 | 807,553 |
| *Obsessive-compulsive disorder* | European ancestry individuals from the International Obsessive Compulsive Disorder Foundation Genetics Collaborative and OCD Collaborative Genetics Association Studies | 2,688 | 7,037 | 9,725 |
| *Schizophrenia* | Ninety cohorts including ~80% European ancestry participants and ~20% East Asian ancestry participants | 69,369 | 236,642 | 306,011 |
| *Educational attainment<sup>†</sup>* | Sixty-nine European ancestry cohorts in the Social Science Genetic Association Consortium and the UK Biobank European ancestry individuals |  |  | 765,283 |
| *Cognitive performance<sup>‡</sup>* | Twenty-four European ancestry cohorts in the Cognitive Genomics Consortium and the UK Biobank European ancestry individuals |  |  | 257,828 |

<sup>†</sup>Educational attainment was measured as number of years of schooling completed
<sup>‡</sup>Cognitive performance was quantified based on various tests conducted in each study cohort

### Power calculation for Mendelian randomization

To assess the likelihood of type II errors, we conducted power calculations for MR analyses using the mRnd R package^43^ (https://shiny.cnsgenomics.com/mRnd/, accessed December 15, 2023). The type I error rate was set at 5%. For each psychiatric disorder, the statistical power was derived by assuming a true odds ratio (OR) per one standard deviation change in vitamin B12 levels of 1.01, 1.02, 1.03, 1.04, 1.05, 1.10, or 1.20, respectively. For educational attainment and cognitive performance, the statistical power was derived by assuming a true association with a magnitude of 0.01, 0.02, 0.03, 0.04, 0.05, 0.10, or 0.20 per one standard deviation change in vitamin B12 levels, respectively.

### Associations between vitamin B12 levels and psychiatric disorders, educational attainment, and cognitive performance

Similar to positive control analyses, we performed MR to assess the association between the genetically predicted vitamin B12 levels and each of the outcomes. MR estimates derived using the weighted median method were reported as primary results. Associations with a p-value < 5.0x10^-3^ (Bonferroni-corrected significance threshold accounting for 10 outcomes) were considered significant. Secondary analyses were performed using the inverse variance weighted regression, penalized weighted median, simple mode, weighted mode, and MR-Egger method, respectively. For folate levels, since two genetic instruments were used, the inverse variance weighted regression estimates were derived to assess the association with each outcome.

### Sensitivity analyses excluding pleiotropic genetic instruments

To mitigate potential bias from horizontal pleiotropy, we excluded genetic instruments that were subject to a high risk of horizontal pleiotropy and repeated the MR analyses for vitamin B12 levels. Subsequently, we further excluded genetic instruments that were subject to a moderate risk of horizontal pleiotropy and again repeated the MR analyses for vitamin B12 levels.

### Within-sibship genome-wide association studies

Since population GWAS estimates for psychiatric disorders, educational attainment, and cognitive performance may be confounded by uncontrolled demographic and indirect genetic effects, we repeated the MR analyses leveraging within-sibship GWAS estimates that were available for depressive symptoms, educational attainment, and cognitive performance^44^. Notably, unlike in the GWAS for depression, depressive symptoms were measured using various rating scales and standardized to be a continuous outcome^44^. European ancestry-specific within-sibship GWAS summary statistics were obtained from the Within Family Consortium^44^. Details of these GWASs have been described previously and are summarized in **Supplementary Table S3**.

## Results

### Mendelian randomization confirmed association between vitamin B12 levels and pernicious anemia

An overview of this study is illustrated in **Figure 1**. Meta-analyses of GWASs for serum vitamin B12 levels identified 11 genetic instruments (p-value < 2.2x10^-9^; **Supplementary Table S1**). Interestingly, all of these 11 variants are missense or stop-gain variants of protein-coding genes (**Supplementary Table S1**). Of these genetic instruments, the minimal F-statistic was 43.14, suggesting a low risk of weak instrument bias (**Supplementary Table S1**). Notably, some of these genetic instruments may pose a risk of horizontal pleiotropy. For instance, rs602662, a missense variant of *FUT2*, was subject to a high risk of horizontal pleiotropy (**Supplementary Table S2**). Missense variants in *MUT*, *FUT6*, *CD320*, and *CUBN* were subject to a moderate risk of horizontal pleiotropy due to their associations with other phenotypes, such as liver function biomarkers, lipid levels, and height (**Supplementary Table S2**).

**Figure 1.**
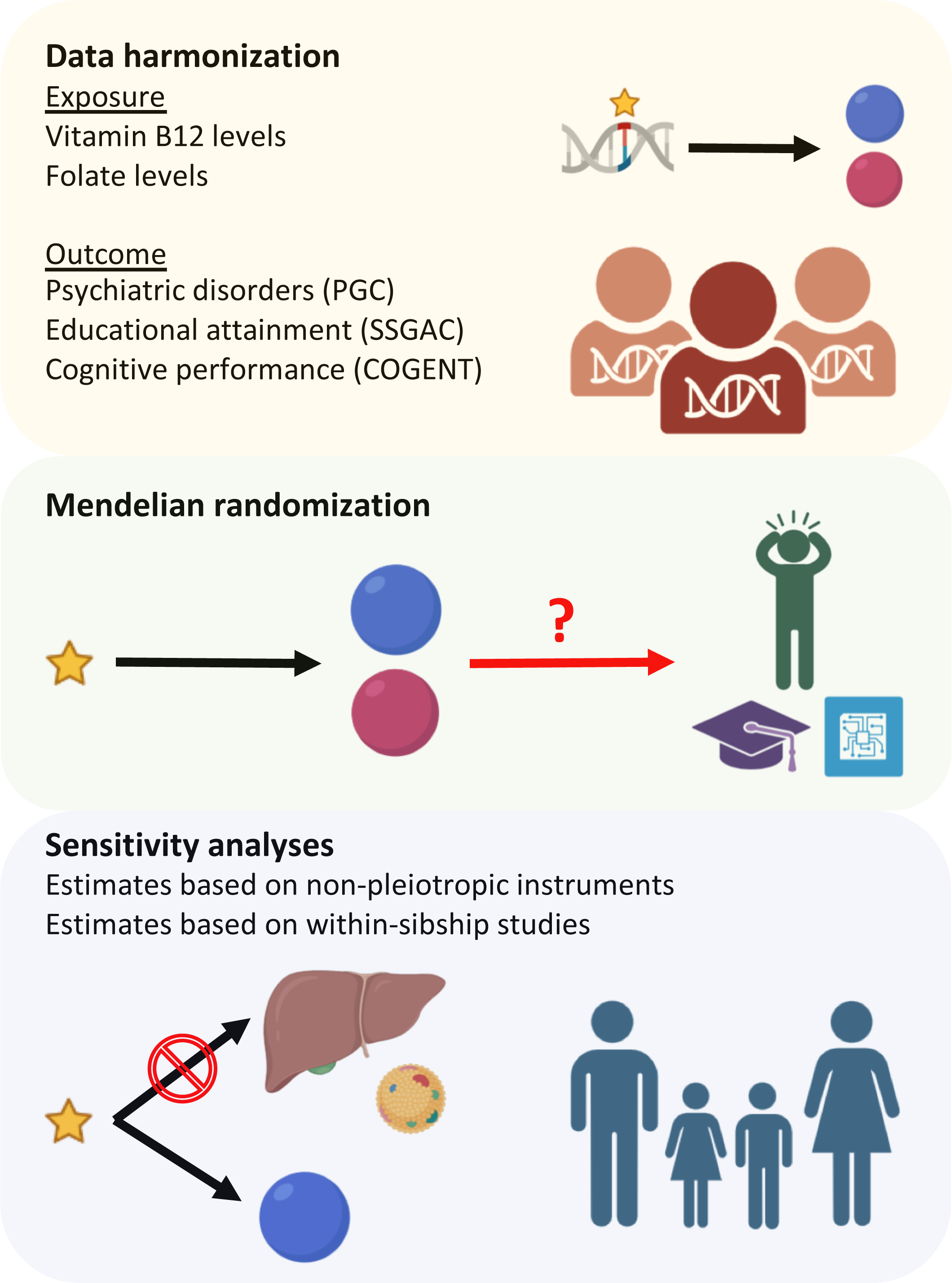
Overview of study design. Summary statistics of large-scale genome-wide association studies were obtained for serum vitamin B12 and folate levels, eight psychiatric disorders, educational attainment, and cognitive performance. Mendelian randomization and multiple sensitivity analyses were performed to estimate the potential effects of vitamin B12 and folate levels.

Based on the UK Biobank, we confirmed that a one standard deviation increase in genetically predicted vitamin B12 levels was strongly associated with an OR of 0.27 for vitamin B12 deficiency anemia based on ICD-10 codes (95% CI: 0.19-0.40; p-value = 1.0x10^-11^), as well as an OR of 0.24 for self-reported pernicious anemia (95% CI: 0.15-0.40; p-value = 2.1x10^-8^; **Figure 2** and **Supplementary Table S4**). Meanwhile, the genetically predicted vitamin B12 levels were not associated with other types of anemia (**Figure 2** and **Supplementary Table S4**). MR estimates derived using different methods were highly consistent (**Supplementary Table S4**).

**Figure 2.**
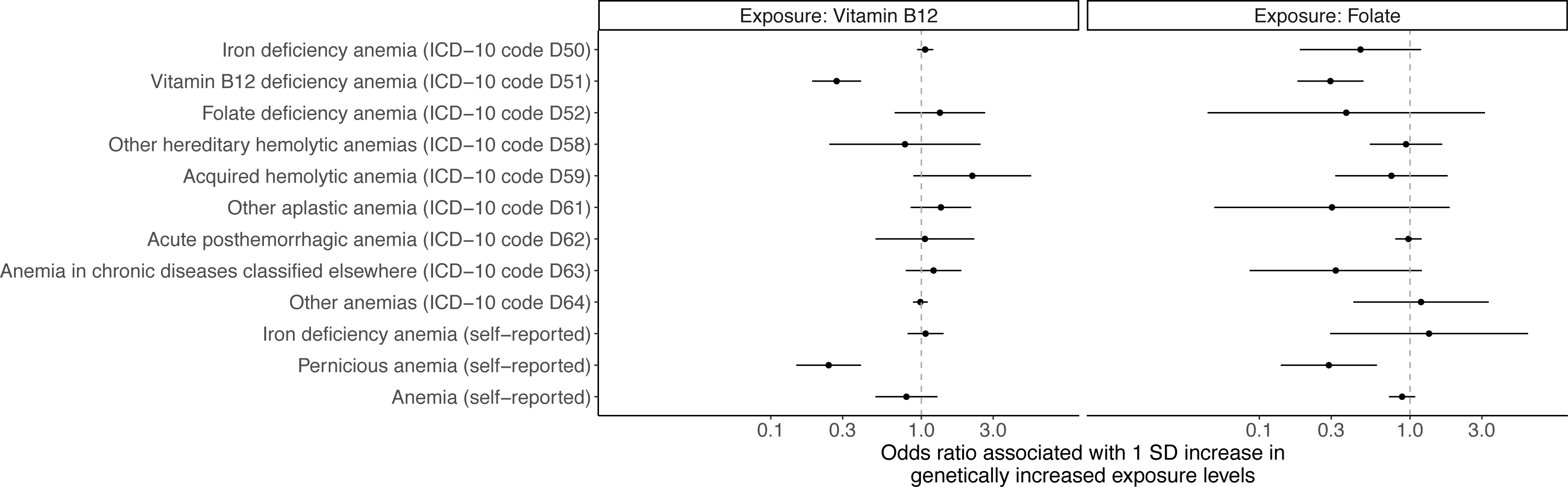
Associations between genetically predicted vitamin B12 and folate levels and various types of anemia. Mendelian randomization estimates obtained using the weighted median method and the Wald ratio method are illustrated for vitamin B12 levels and folate levels, respectively.

In addition, two independent genetic variants, rs1801133 and rs652197, were significantly associated with serum folate levels (p-value < 2.2x10^-9^; **Supplementary Table S1**). However, rs1801133, a missense variant of *MTHFR*, demonstrated a high risk of horizontal pleiotropy, while rs652197, an intronic variant of *FOLR3*, has been associated with serum 25-hydroxyvitamin D levels in previous studies (**Supplementary Table S2**). As expected, the genetically predicted folate levels were also associated with a reduced odds of vitamin B12 deficiency anemia based on ICD- 10 codes (OR = 0.30; 95% CI: 0.18-0.49; p-value = 2.3x10^-6^) and self-reported pernicious anemia (OR = 0.29; 95% CI: 0.14-0.60; p-value = 9.3x10^-4^; **Figure 2** and **Supplementary Table S4**). Although folate levels were predicted to reduce the odds of folate deficiency anemia, this association was not significant, likely due to the small number of folate deficiency anemia cases (**Figure 2** and **Supplementary Table S4**).

### Genetically predicted vitamin B12 levels were not associated with psychiatric disorders, educational attainment, and cognitive performance

The large-scale GWASs for psychiatric disorders, educational attainment, and cognitive performance ensured sufficient statistical power in MR analyses (**Table 1** and **Supplementary Table S5**). Of the eight psychiatric disorders under investigation, the GWAS meta-analyses for depression had the largest sample size. As a result, with a type I error rate of 5%, MR analyses with depression as the outcome could achieve a power ≥ 80% when the true OR per one standard deviation change in vitamin B12 levels was equal to or greater than 1.03 (**Supplementary Table S5**). Comparably, MR analyses for educational attainment and cognitive performance could achieve a power of 100% and 94%, respectively, when the true association had a magnitude of 0.03 per one standard deviation change in vitamin B12 levels (**Supplementary Table S5**).

Despite sufficient statistical power, MR analyses did not detect any significant associations between genetically predicted vitamin B12 levels and any outcomes (**Figure 3** and **Supplementary Table S5**). For example, a one standard deviation increase in genetically predicted vitamin B12 levels was associated with an OR of 1.01 for depression (95% CI: 0.97-1.04; p-value = 0.74), as well as a 7.7x10^-3^ and 1.3x10^-2^ standard deviation increase in educational attainment (95% CI: -1.0x10^-2^-2.5x10^-2^; p-value = 0.39) and cognitive performance (95% CI: - 8.8x10^-3^-3.5x10^-2^; p-value = 0.24), respectively (**Figure 3** and **Supplementary Table S5**). The estimated associations between folate levels and the outcomes also largely overlapped with the null (**Figure 3** and **Supplementary Table S6**).

**Figure 3.**
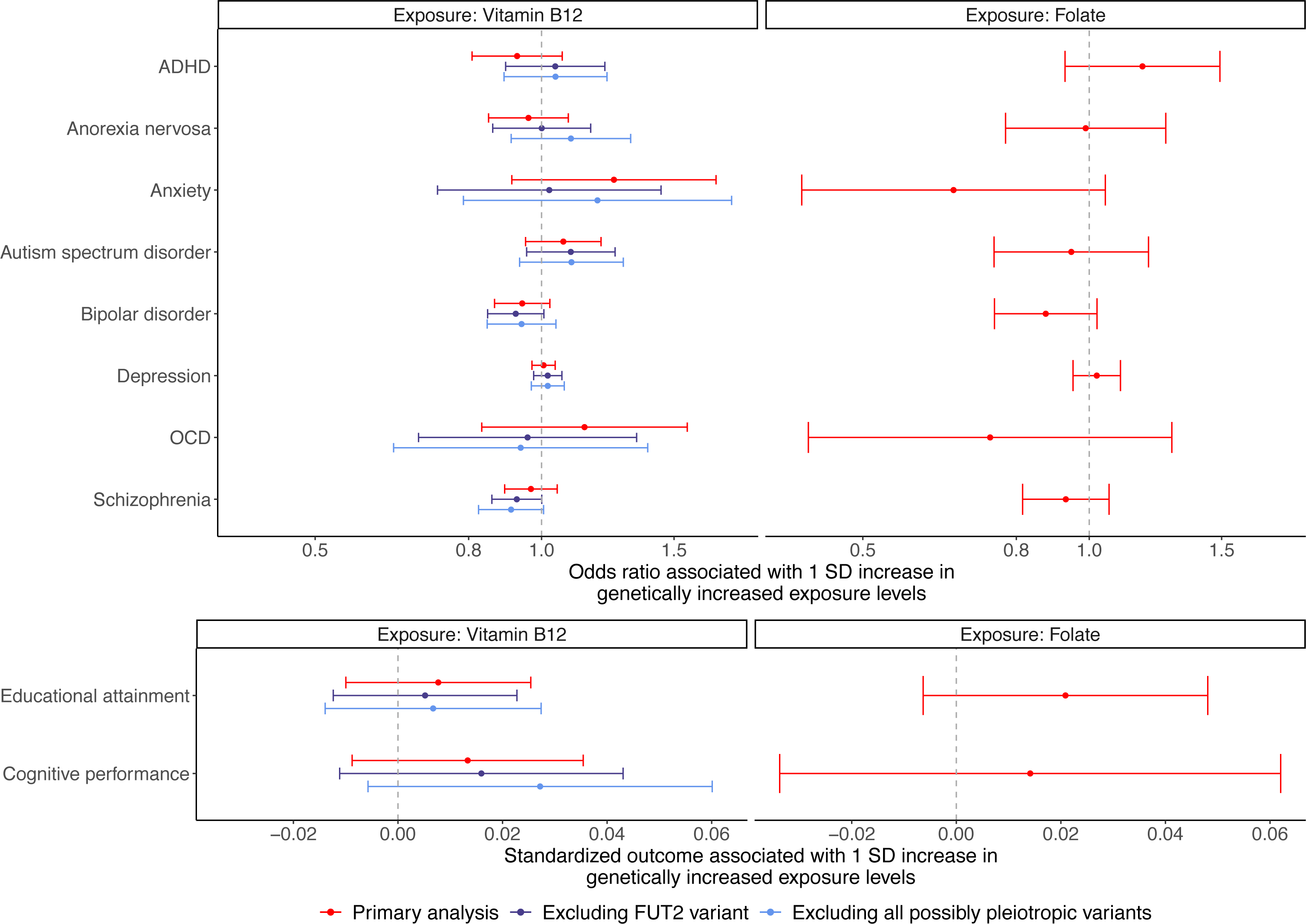
Associations between genetically predicted vitamin B12 and folate levels and eight psychiatric disorders, educational attainment, and cognitive performance. Sensitivity analyses for vitamin B12 levels were performed after excluding genetic instruments that were potentially subject to horizontal pleiotropy. Mendelian randomization estimates obtained using the weighted median method and the inverse variance weighted regression method are illustrated for vitamin B12 levels and folate levels, respectively.

Furthermore, no associations were detected between genetically predicted vitamin B12 levels and any outcomes in sensitivity analyses where the genetic instruments demonstrating potential horizontal pleiotropy were excluded (**Figure 3** and **Supplementary Tables S7** and **S8**). In fact, none of the tested associations had a nominal p-value < 0.05 in the primary analyses using the weighted median method (**Figure 3**). The estimates derived using alternative methods were highly consistent with the primary results, with the only exception that with the penalized weighted median method, a one standard deviation increase in genetically predicted vitamin B12 levels was associated with an OR of 0.82 for ADHD (95% CI: 0.72-0.93; p-value = 2.1x10^-3^; **Supplementary Figure S1** and **Supplementary Table S5**). Nevertheless, this association was not detected after excluding the highly pleiotropic genetic instrument in *FUT2* (**Supplementary Table S7**) as well as after further excluding the moderately pleiotropic genetic instruments (**Supplementary Table S8**).

### Sensitivity analyses based on within-sibship genome-wide association studies

MR analyses leveraging within-sibship GWASs for depressive symptoms, educational attainment, and cognitive performance did not identify any significant associations with genetically predicted vitamin B12 levels (**Figure 4A**). Specifically, a one standard deviation increase in genetically predicted vitamin B12 levels was associated with a 5.4x10^-2^ standard deviation decrease in depressive symptoms (95% CI: -0.14-3.6x10^-2^; p-value = 0.24), as well as a 5.6x10^-3^ and 3.9x10^-2^ standard deviation increase in educational attainment (95% CI: -4.3x10^-2^-5.5x10^-2^; p-value = 0.82) and cognitive performance (95% CI: -6.6x10^-2^-0.14; p-value = 0.47), respectively (**Figure 4A** and **Supplementary Table S9**). These estimates were consistent with those obtained using alternative MR methods (**Figure 4B-D** and **Supplementary Table S9**).

**Figure 4.**
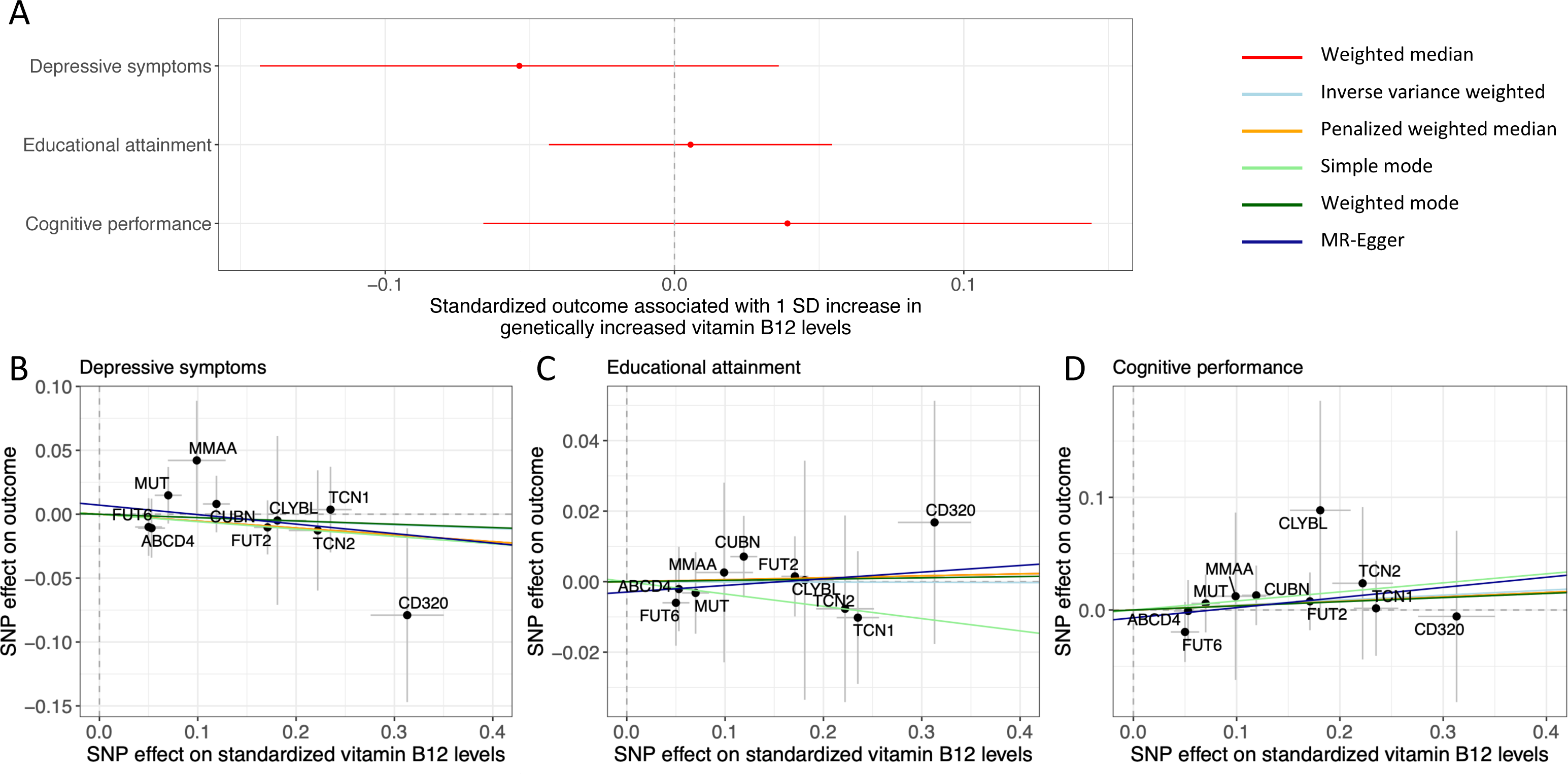
Estimated effects of vitamin B12 levels based on within-sibship studies. (A) Associations between genetically predicted vitamin B12 and depressive symptoms, educational attainment, and cognitive performance. Mendelian randomization estimates obtained using the weighted median method are illustrated. Scatter plots comparing the genetic instrument-outcome and the genetic instrument-vitamin B12 levels associations are illustrated for (B) depressive symptoms, (C) educational attainment, and (D) cognitive performance. The mapped genes of the respective genetic instruments are indicated. The slopes of the colored lines represent estimates obtained through different Mendelian randomization methods.

## Discussion

Vitamin B12 has important roles in one-carbon metabolism as both an enzyme cofactor or substrate^1^. Individuals with vitamin B12 deficiency can develop pernicious anemia as well as neuropsychiatric diseases^7, 8^. However, it remains unclear whether the general population may benefit from vitamin B12 supplementation for protection against psychiatric disorders and cognitive impairment. In this study, we performed MR analyses to estimate the potential effects of serum vitamin B12 levels on eight psychiatric disorders, educational attainment, and cognitive performance. We did not detect any significant associations between genetically predicted serum vitamin B12 levels and any of these outcomes.

Our study has several strengths. First, we employed genetic instruments that were protein-altering variants affecting crucial genes in the absorption, transport, or enzymatic reactions of vitamin B12 for MR analyses^18^. This approach effectively mitigated bias arising from unmeasured confounding and reverse causation. Second, the use of large-scale GWASs for the outcomes, particularly depression, educational attainment, and cognitive performance, minimized the likelihood of type II errors, given the reporting of null associations. These results were compared to the positive control, where genetically increased vitamin B12 levels exhibited strong protective effects against pernicious anemia, thereby supporting the validity of our analyses. Third, consistent MR estimates were obtained in sensitivity analyses where pleiotropic variants were removed, safeguarding an indispensable instrumental variable assumption of MR. Nevertheless, MR estimates for folate levels were more prone to bias due to horizontal pleiotropy, thus interpreting the results involving folate levels requires extra caution. Fourth, MR analyses based on within-sibship GWASs for depressive symptoms, educational attainment, and cognitive performance further reduced the risk of confounding due to uncontrolled population stratification, assortative mating, or indirect genetic effects^44^.

Our study has a clear and important implication that, in the general population, vitamin B12 supplementation is unlikely to meaningfully reduce the risks of the investigated psychiatric disorders or significantly improve educational attainment or cognitive performance. These results may discourage randomized placebo-controlled trials amongst individuals without clinical vitamin B12 deficiency, while encouraging the identification of other risk factors and preventive measures for psychiatric disorders and cognitive impairment. However, it is noteworthy that none of the GWASs utilized in this study were based on populations selected for vitamin B12 deficiency. Multiple lines of evidence are still needed to ascertain the potential impact of vitamin B12 or folate plus vitamin B12 supplementation on various outcomes in individuals with vitamin B12 deficiency.

Some limitations of our study should be noted. First, our MR analyses relied on sex-combined GWASs and could not identify potential sex-specific effects of vitamin B12 levels, since large-scale sex-stratified GWASs for both vitamin B12 levels and the outcomes are yet to emerge. Second, our analyses were restricted to populations of European ancestry. It remains unclear whether our findings could be generalized to other populations of non-European ancestries. Third, larger GWASs for certain psychiatric disorders, such as OCD and anxiety disorders, are necessary to increase the statistical power of MR analyses for identifying potential effects of small magnitudes.

Fourth, our MR analyses could estimate population-averaged associations between vitamin B12 levels and the outcomes, but not potential dose-dependent effects of vitamin B12 levels, which would require availability of both vitamin B12 measurements and the outcomes in the same study population^45, 46^. Fifth, one of the genetic instruments, rs371753672, a missense variant of *MMACHC*, was not available in any GWASs utilized in this study due to its low minor allele frequency. However, this variant only captures a small proportion of variance in vitamin B12 levels. Last, our results do not provide information on whether serum vitamin B12 levels may influence the progression of psychiatric disorders or cognitive impairment, as the genetic architecture underlying disease progression may differ from that underlying disease pathogenesis. We anticipate that future investigations will more comprehensively illuminate the role of vitamin B12 in various psychiatric disorders and cognitive impairment in diverse populations.

In conclusion, through MR analyses, we demonstrated that genetically predicted serum vitamin B12 levels were not associated with eight psychiatric disorders, educational attainment, or cognitive performance. Our findings indicate that vitamin B12 supplementation is unlikely to offer protection against these psychiatric disorders or cognitive impairment in the general population.

## Author contributions

T.L. and A.D.P. conceptualized the study. T.L. curated the data, performed the analyses, and wrote the original draft. T.L. and A.D.P. interpreted the results, and reviewed and edited the manuscript critically.

## Data Availability

All data produced in the present work are contained in the manuscript

## Acknowledgements

We thank David S. Rosenblatt and David Watkins for helpful discussions. T.L. has been supported by a Schmidt AI in Science Postdoctoral Fellowship. The funders have no role in study design; collection, management, analysis and interpretation of data; or the decision to submit for publication.

---

**Figure S1.**
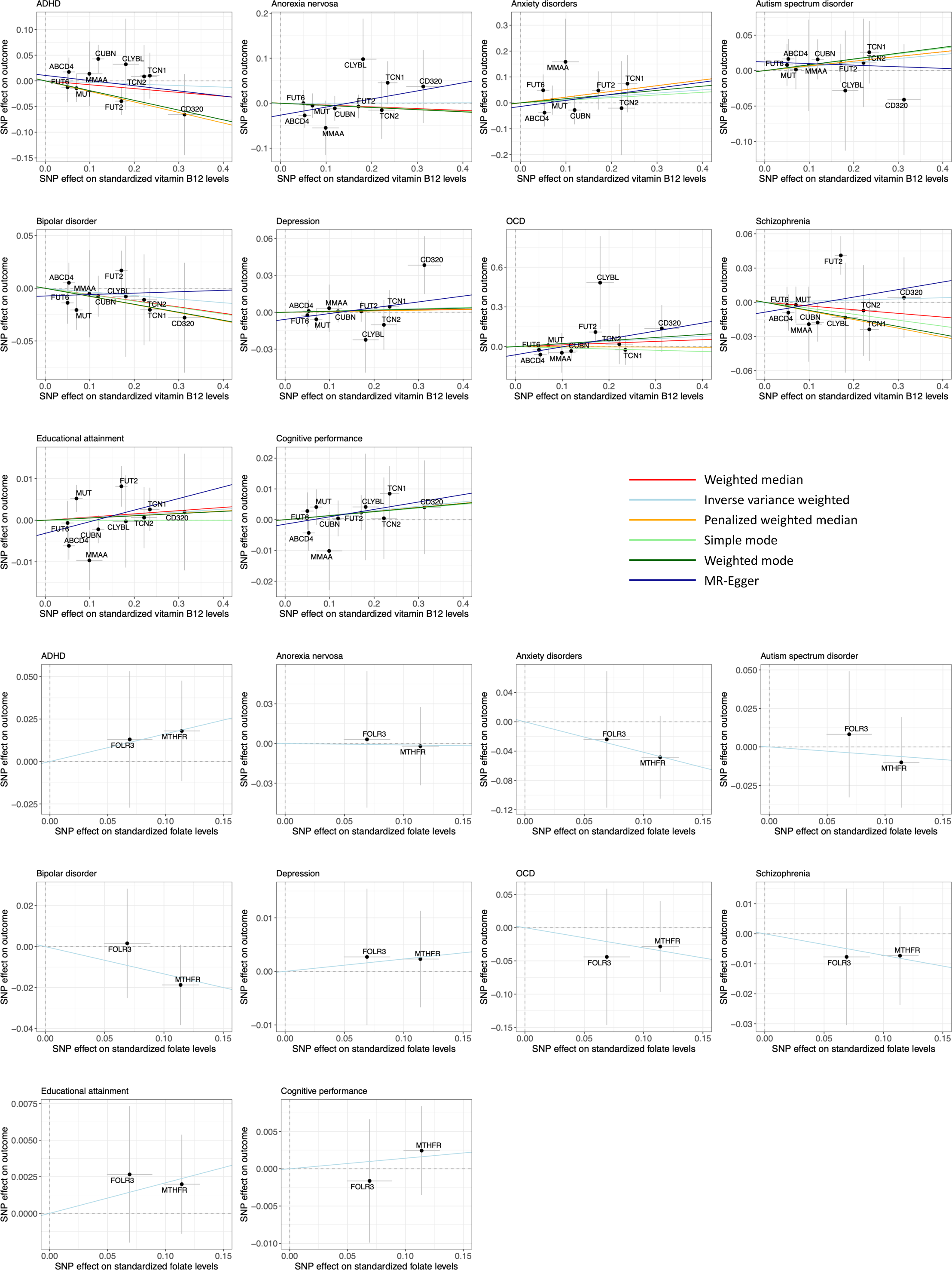
Scatter plots comparing the genetic instrument-outcome and the genetic instrument-exposure associations. Primary results were obtained using the weighted median method and the inverse variance weighted regression method for vitamin B12 levels and folate levels, respectively. The mapped genes of the respective genetic instruments are indicated. The slopes of the colored lines represent estimates obtained through different Mendelian randomization methods.

